# Ancestry specific distribution of LPA Kringle IV-Type-2 genetic variants highlight associations to apo(a) copy number, glucose, and hypertension

**DOI:** 10.1101/2024.07.09.24310176

**Authors:** Yihao Li, Florian Kronenberg, Stefan Coassin, Badri Vardarajan, Gissette Reyes-Soffer

**Affiliations:** Gertrude H. Sergievsky Center, Dept of Neurology, Columbia University Vagelos College of Physicians and Surgeons, 630 West 168th Street, PH19-306, New York, N.Y.10032; Institute of Genetic Epidemiology, Medical University of Innsbruck, Innsbruck, Austria; Columbia University Vagelos College of Physicians and Surgeons, Department of Medicine, Division of Preventive Medicine and Nutrition, P&S 10-501,New York, NY, USA

**Author notes:** **Corresponding Authors:** Gissette Reyes-Soffer, MD Assistant Professor of Medicine, Columbia University Vagelos College of Physicians and Surgeons, 630 West 168^th^ Street, P&S-10-501, New York, N.Y.10032., Badri Vardarajan, PhD, Gertrude H. Sergievsky Center, Dept of Neurology, Columbia, University Vagelos College of Physicians and Surgeons, 630 West 168^th^ Street, PH19-306, New York, N.Y.10032.

## Abstract

**Background:** High Lp(a) levels contribute to atherosclerotic cardiovascular disease and are tightly regulated by the *LPA* gene. Lp(a) levels have an inverse correlation with *LPA* Kringle IV Type-2 (KIV-2) copy number (CN). Black (B) and Hispanic (H) individuals exhibit higher levels of Lp(a), and rates of CVD compared to non-Hispanic Whites (NHW). Therefore, we investigated genetic variations in the *LPA* KIV-2 region across three ancestries and their associations with metabolic risk factors.

**Methods:** Using published pipelines, we analyzed a multi-ethnic whole exome dataset comprising 3,817 participants from the Washington Heights and Inwood Columbia Aging Project (WHICAP): 886 [NHW (23%), 1,811 Caribbean (C) H (47%), and 1,120 B individuals (29%). Rare and common variants (alternative allele carrier frequency, CF < 0.01 or > 0.99 and 0.01 < CF < 0.99, respectively) were identified and KIV-2 CN estimated. The associations of variants and CN with history of heart disease, hypertension (HTN), stroke, lipid levels and clinical diagnosis of Alzheimer’s disease (AD) was assessed. A small pilot provided in-silico validation of study findings.

**Results:** We report 1421 variants in the *LPA* KIV-2 repeat region, comprising 267 exonic and 1154 intronic variants. 61.4% of the exonic variants have not been previously described. Three novel exonic variants significantly increase the risk of HTN across all ethnic groups: 4785-C/A (frequency = 78%, odds ratio [OR] = 1.45, p = 0.032), 727-T/C (frequency = 96%, OR = 2.11, p = 0.032), and 723-A/G (frequency = 96%, OR = 1.97, p = 0.038). Additionally, six intronic variants showed associations with HTN: 166-G/A, 387-G/C, 402-G/A, 4527-A/T, 4541-G/A, and 4653-A/T. One intronic variant, 412-C/T, was associated with decreased blood glucose levels (frequency = 72%, β = −14.52, p = 0.02).

Three of the associations were not affected after adjusting for *LPA* KIV-2 CN: 412-C/T (β = −14.2, p = 0.03), 166-G/A (OR = 1.41, p = 0.05), and 387-G/C (OR = 1.40, p = 0.05). KIV CN itself was significantly associated with 314 variants and was negatively correlated with plasma total cholesterol levels.

**Conclusions:** In three ancestry groups, we identify novel rare and common *LPA* KIV-2 region variants. We report new associations of variants with HTN and Glucose levels. These results underscore the genetic complexity of the *LPA* KIV-2 region in influencing cardiovascular and metabolic health, suggesting potential genetic regulation of pathways that can be studied for research and therapeutic interventions.

**Clinical Perspective:**

- Lp(a) levels are mostly controlled by the *LPA* gene and are higher in Blacks and Hispanics.
- *Novel LPA* KIV-2 variants found in three ancestry groups, including data on Caribbean Hispanics, show strong positive associations to hypertension and negative associations to glucose levels.
- Further characterization of these variants and identifying links to disease can help precision medicine efforts to understand disease mechanisms in all populations.

## Introduction

Lipoprotein(a) [Lp(a)] is an apolipoprotein B100 (apoB100) containing lipoprotein, with an additional protein component - apolipoprotein little (a) [apo(a)]. Both genetic [through the *LPA* gene locus that codes for apo(a)] and non-genetic factors influence Lp(a) concentrations ^1, 2^ and high levels are linked to increases in atherosclerotic cardiovascular disease (ASCVD) and mortality. The strong genetic regulation by the *LPA* gene allowed the use of genome wide population studies which support its role as a causal risk factor of atherosclerotic cardiovascular disease (ASCVD) ^3, 4^, independent of classical risk factors including other plasma lipids, including low density lipoprotein (LDL).

The *LPA* gene evolved from the plasminogen (*PLG*) gene^5^, the lack of expression in murine models has limited basic research advances that can translate to humans. Significant differences exist in the *LPA* gene compared to *PLG*, one being the expansion of a highly polymorphic region – the Kringle (K)IV Type 2 (KIV-2)^4^. KIV-2 is encoded by a 5.6 kb-sized region that contains 2 exons (sized 160 and 182 bp) spaced by a 4 kb-sized intron. And KIV-2’s copy number (CN) variation determines the CN of KIV repeat with all the other KIV region without CN variation, namely KIV-1 and KIV-3 to KIV-10, considered. The KIV repeat is transcribed into mRNA and translated into apo(a) isoform proteins. The size of isoforms is determined by the CN of the KIV repeat and more than 40 protein expressing isoforms have been described, with most individuals expressing two isoforms and one, generally the smaller size isoform, being dominant^6^. Individuals expressing a low number of KIV repeats produce small apo(a) isoforms (up to 22 KIV repeats) and this generally associates with higher Lp(a) concentrations, whereas individuals carrying large apo(a) isoforms (>22 KIV repeats) have lower Lp(a) concentrations^7^. Mendelian randomization and large meta-analysis studies showed that small isoforms are linked to increased risk of cardiovascular disease^6^. However, the regulation of isoform expression is not well understood^8^.

Several genetic variants that affect Lp(a) plasma levels have been reported and recently reviewed^8^. Importantly, the frequencies for these genetic variants vary by ancestry^9–11^. Lastly, these genetic analysis, have been used to establish high levels of Lp(a) as a top driver for aortic valve stenosis^12^, peripheral arterial disease (PAD)^13^, an inverse relationship between levels and Type 2 diabetes mellitus (T2DM) and more recently have questioned a “genetic” based causal association between Lp(a) levels and venous thrombosis^14, 15^. Although these results have assisted with further characterizing the roles of Lp(a) in disease, the translation linking Lp(a) levels and isoforms to direct clinical outcomes needs further validation.

Recent research has highlighted the importance of the *LPA* gene region, particularly in analyzing single nucleotide variants (SNVs) and their relationship with the copy number (CN) of KIV repeats and cardiovascular disease (CVD)^16^. Given the disparities noted in KIV variants across different populations and the limited data available from diverse cohorts, coupled with the heightened risk and elevated Lp(a) levels observed in Black and Hispanic populations, we utilized a recently published protocol ^17^ to identify both common and rare variants within the KIV repeat region of the *LPA* gene. In this study, we present ancestry-specific frequencies for Non-Hispanic White (NHW), Caribbean Hispanic (CH), and Black individuals (B), and explore their associations with heart disease, lipid levels, stroke, Alzheimer’s Disease (AD), and Type 2 Diabetes Mellitus (T2DM). Additionally, we investigate how these variants correlate with estimated KIV CN.

## Methods

### Study Population

We analyzed existing data from 3817 individuals enrolled in the Washington Heights/Inwood Columbia Aging Project (WHICAP), encompassing individuals with various disease conditions and healthy controls from three ancestry groups. This cohort had not been previously assessed for lipid phenotypes. Within the cohort there are genetically identified NHW (23%), African Americans (29%) [we will refer to this cohort as (B) individuals], CH (47%) from the Dominican Republic and Puerto Rico, and 68% are female. Sex is recorded as self-identified. The use of the WHICAP cohort (#AAAO9804) and additional studies presented here (#AAAV0773) are approved by the institutional review board of Columbia University, New York City. All participants provided written informed consent. The WHICAP stored data included standardized medical and neurological examinations, antecedent risk factors, cognitive assessments, genome wide association study (GWAS) for AD, whole exome (WES), whole genome sequencing (WGS) and fasting plasma lipid levels. The details of the WES and data processing are described here^18^. The study also has stored serum and plasma samples that can be used, provided consent for future research was obtained. The latter allowed for the measurement of Lp(a) levels and isoform size in our pilot study.

### Identification of *LPA* KIV-2 variants by ancestry

*LPA* KIV-2 Variant calling in WES: Previous studies have estimated that the *LPA* gene can explain 70-90% of the variance of Lp(a) plasma protein levels. As previously mentioned, the gene contains a large CN variation that expresses over 40 protein isoforms^15^. This variable number of repeats causes large apo(a) size polymorphism. Because this region is hyper-variable and difficult to map using standard sequence analysis protocols, variants within the KIV-2 region were usually missed in common sequencing projects, leaving up to 70% of the *LPA* coding region unaddressed. We used a recently published methodology ^17^ to analyze this region in the WHICAP cohort.

The WHICAP whole exome sequencing (WES) data set has been aligned to human genome b37 using the BWA-mem pipeline as described before^18^. We used a specialized pipeline to determine the genetic variation in the *LPA* gene as described here^17^. Briefly, for each participant, we aligned exome-wide reads to the *LPA* KIV-2 (6^th^ repeat) reference sequence^19^ (i.e. hg19 6:161,033,785-161,038,888) assembled using an ultra-deep sequencing protocol. Following realignment to *LPA* KIV-2, reads with high base, mapping, and alignment quality metrics, we called high quality variants using a method originally developed for detecting mitochondrial DNA heteroplasmy^8^. For each variant, the pipeline produces a present/absent call but the information about genotype dosage is not provided. The identified variants were annotated with the location of genetic elements (exons, introns, etc.) and exon reading frame. Variants were excluded/dropped if they: (a) were missing in more than 10% of individuals, (b) the individual in the cohort had a total coverage < 50 reads at this variant or (c) variant was monomorphic. Further, we defined variants found in >1% of the participants as common polymorphisms. The resulting variant profiles for all WHICAP participants are reported.

### WHICAP cohort data: use of existing data

In 3817 individuals, we tested the association of *LPA* KIV-2 variants, identified from the pipeline described above, with cardio and cerebrovascular phenotypes including history of heart disease, stroke, hypertension (HTN), clinical diagnosis of AD, lipid levels [total cholesterol (TC), calculated LDL-C, TG, HDL](mg/dL), self-reported diagnosis of T2DM, glucose levels(mg/dL), hemoglobin (HbA1C), Insulin(MIU/ml) and C-Peptide(ng/dL). The availability of these outcomes was limited to a subset individuals as shown in **Table 1**. Details and validation of the self-reported variables capturing history of heart disease and stroke are provided elsewhere^20^. Diagnosis of clinical AD was established based on the NINCDS-ADRDA criteria^21,22^. Description of methods used to obtain outcome data in WHICAP including assay for measurements have been previously described ^23^.

**Table 1.** Study population and baseline characteristics. **Legend**: Baseline characteristics for study cohort. *The sex for two participants was missing (only 3815 individuals are accounted for this phenotype). ** Lipid data available in 1011 individuals. N= Available data set. NHW: non-Hispanic White, B: Blacks, CH: Caribbean Hispanic. AD: Alzheimer’s Disease, T2DM: type 2 diabetes mellitus. Clinical diagnosis (AD, History of heart disease, DM, Stroke and HTN) were not available for the full cohort. LDL-C: low density lipoprotein cholesterol; HDL: high density lipoprotein. ± represents mean and standard deviation for normally distributed variables; ( ) represents median and interquartile range. One outlier in Insulin was further dropped (extremely out of range level).

| Characteristic | Study Population |  |
| --- | --- | --- |
| Age, y | $80.6 \pm 7$ | |
| Age range | 62-109 |  |
| Full cohort race | Female -N = 2606 (68.3%)* | Male(N = 1209 (31.7%)* |
| NHW | 521(13.66%) | 364(9.54%) |
| AA | 793(20.79%) | 327(8.57%) |
| CH | 1292(33.87%) | 518(13.58%) |
| Clinical characteristics | Number of cases (%) |  |
| Clinical AD, N=3755 | 1747 (46.52%) |  |
| History of Heart Disease, N=3815 | 1477(38.72%) |  |
| Diabetes mellitus, N=3815 | 1137/3815(29.80%) |  |
| Stroke, N=1658 | 103/1658(6.21%) |  |
| Hypertension, N=3815 | 3114/3815(81.63%) |  |
| Race N=1011** | Female - N = 695 (68.7%) | Male - N = 316 (31.3%) |
| NHW | 183(18.10%) | 126(12.46%) |
| AA | 220(21.76%) | 87(8.61%) |
| CH | 292(28.88%) | 103(10.19%) |
| Total cholesterol, mg/dL | $197.7 \pm 38.4$, N = 1011 | |
| Triglycerides, mg/dL | 131.0 (93-182), N = 1011 |  |
| LDL-C, mg/dL | $99.1 \pm 32.9$, N = 830 | |
| HDL-C, mg/dL | $47.6 \pm 14.1$, N = 1011 | |
| Insulin, MIU/mL | 14.8 (7.4-30.8), N = 836 |  |
| Glucose, mg/dL | $113.9 \pm 48.1$, N = 837 | |
| Hemoglobin A1C, % | $6.3 \pm 0.9$, N = 464 | |
| C-Peptide, ng/dL | 3.63 (2.3-5.9), N = 836 |  |

Associations were tested using independent regression models within each ancestry (ordinary linear for quantitative traits and logistic for binary traits), adjusting for age at blood draw, sex and first three principal components (computed from genome-wide SNP data) to account for population substructure. Insulin, C-Peptide and Lp(a) levels were log transformed as they were not normally distributed (right skewed). All data was analyzed using Statsmodels 0.13.1 running on Python 3.8. The results were summarized using an inverse variance meta-analysis approach implemented in (meta-analysis tool) METAL 11.3.25^24^. Variant association in each ancestry group was included only when at least five individuals carried the minor allele of the specific variant. Statistical significance was established using a false discovery rate (FDR) adjusted p-value = 0.05 threshold^25^. For details of other modules involved, please check the GitHub repository (https://github.com/alexliyihao/lpa-analysis). We detected haplotypes amongst variants that had strong associations to clinical phenotypes using Haploview^26^. Because the LPA variant calling pipeline outputs presence/absence calls for variants and doesn’t genotype those calls, we used 0/1 status (instead of variant dosage) for computing LD metrics (D’ and r^2^).

### Estimation of *LPA* KIV-2 copy number (CN)

We deployed the VNTR calling pipeline as described here^27^ on hg19 reference estimating the diploid length (i.e. the sum of both allele) of *LPA* KIV-2 variable number tandem repeats (VNTR) on the WHICAP cohort. Code and details of other modules involved are shared via GitHub repository (https://github.com/alexliyihao/vntrwrap). This pipeline estimates the read depth of KIV-2 types 1A and 1B and give normalizations from (a) the mean depth extracted for all the regions labelled “extracted” from this individual and (b) 200 closest individuals in the cohort from sequencing profile perspective, then predicts the KIV-2 CN by a scaling factor of 34.9 and 5.2 respectively and an additional adjustment of - 1 to account for the inclusion of invariant from KIV-1 and KIV-3 in the estimation. We tested the association of this estimates with all WHICAP phenotypes and *LPA* KIV-2 variants.

In addition to assessing the copy number (CN) data, previous studies have reported the distribution of KIV-2 types (i.e. A, B, or C) ^8, 17^. In this study, we did not adjust for these subtypes as their effect has not been studied in large diverse cohorts and their alignment using WES data may not provide reliable data.

### Plasma Lp(a) levels and isoform size measurements: Pilot Data

Lp(a) plasma concentration was measured using the isoform-independent sandwich ELISA developed by the Northwest Lipid Metabolism and Diabetes Research Laboratory^28^. Apo(a) isoform size estimates were performed by the same laboratory using gel electrophoresis and has been previously published by our group^29^. We calculated weighted isoform size (*wIS*)^29^ for all individuals and examined the correlation of Lp(a) levels and *wIS* with identified KIV-2 variants and KIV-2 CN estimate, respectively. Correlations were only tested for variants observed in at least five of the 27 participants for whom we obtained plasma Lp(a) and isoform measurements.

### APO E4/E2 genotype phenotype

APOE e4 status was generated either using either isoelectric focussing described by Hixon-Vernier approach^30^ or TaqMan® SNP genotyping assays (Applied Biosystems, ThermoFisher Scientific, USA) of rs429358 (APOE*4) and rs7412 (APOE*2) SNPs. For analysis, we used the dosage of the APOE-ε4 allele (0,1 or 2 alleles per participant).

### Independent data access and analysis

Dr. Reyes-Soffer, corresponding author, has full access to all the data in the study and takes responsibility for its integrity and the data analysis.

### Materials and data availability

The authors have provided supplemental data for all study outcomes and public code repository has been provided (https://github.com/alexliyihao/lpa-analysis, https://github.com/alexliyihao/vntrwrap).

## Results

Whole exome-sequencing (WES) of 3817 WHICAP participants (886 NHW, 1,120 Blacks and, 1,811 CH) were analyzed. The study WES data includes higher number of females (N=2606) compared to males (N=1209) and the mean age of the population is 80.6±7 years. Two individuals did not report sex (hence sex data is presented for 3815 individuals). The age of this data cohort (subset of full WHICAP cohort) is representative of the full WHICAP data set, a population originally enrolled to study risk of AD and dementia. Baseline characteristics of the 3817 study participants are presented on **Table 1**. Additional details on the WHICAP population, whole exome sequencing, and pre-processing have been previously published^18^.

### *LPA* KIV-2 *v*ariants presentation in WHICAP: three ancestry groups

We identified 1421 common and rare variants (**Supplemental Table S1**, excel file). These were present in exonic (267) and intronic (1154) regions of the *LPA* KIV-2 repeat and 61.4% of the exonic variants have not been previously described. **Figure 1** highlights the overlap and presentation of the variants within each ancestral group: 36% of all variants (**Fig 1A**). CH and B had similar percentage of unique variants (20% and 16% respectively- **Fig 1A**). Of the 267 exonic variants we identified, 103 (38.6%) were previously identified (**Figure 1B**) by Coassin et al^8, 17^ while 164 (61.4%) were not previously described (**Figure 1C**). 35% of these novel exonic variants were shared by the three ethnic groups. However, 26% of exonic variants were unique to CH and 21% to B individuals (**Fig 1C**). NHW had only 8% novel variants and this was similar to exonic variant presentation in this group. **Figure 1D** list all novel intronic variants, previous studies have not described intronic variants. **Supplemental Figure S1,** list the locations of the identified variants. Previously reported variants (non-novel) that were found in our cohort and their ancestry appearance frequency compared to the 1000 Genomes Project cohort are listed in **Supplemental Table S2.**

**Figure 1.**
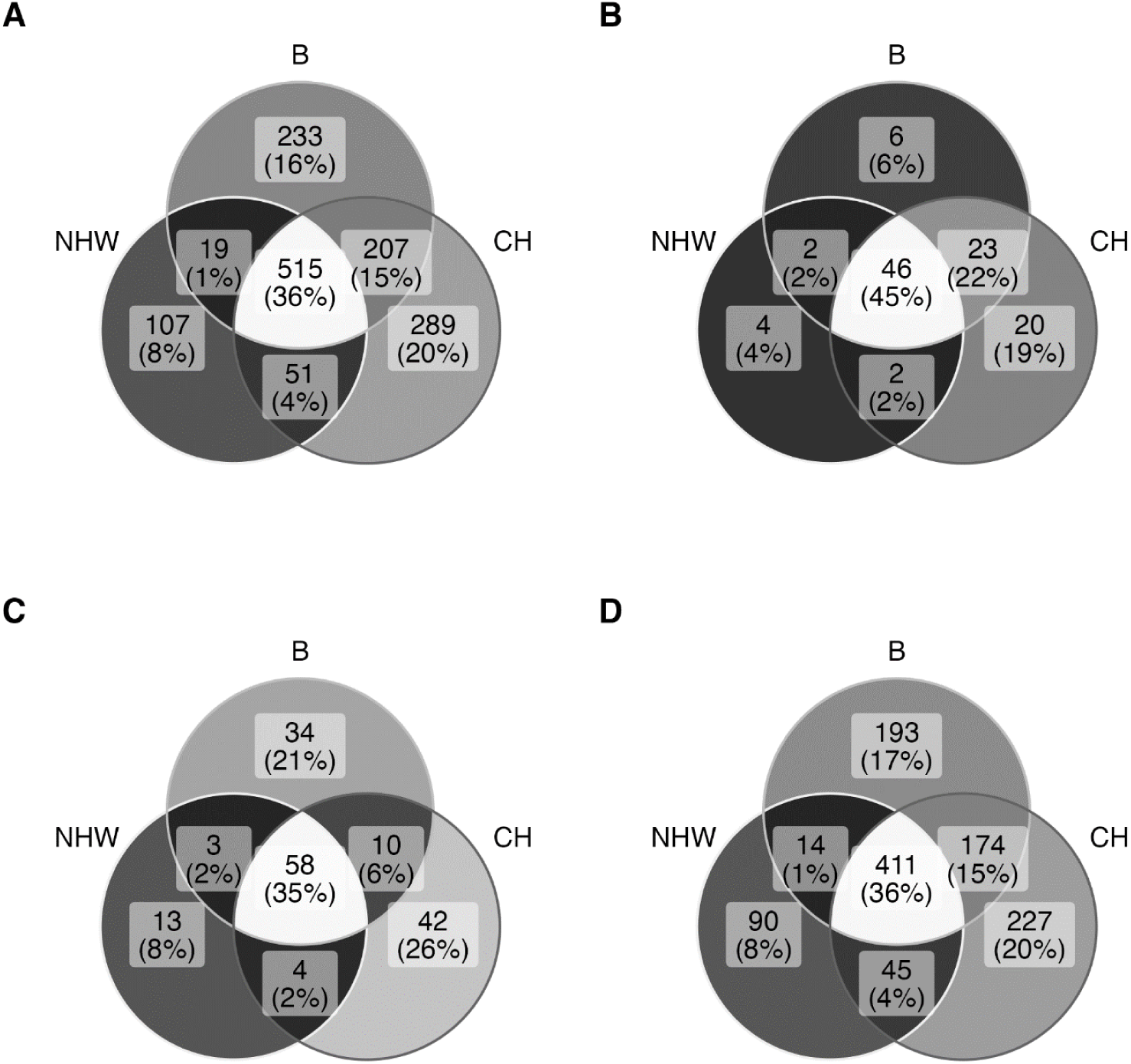
Distribution of WHICAP *LPA* KIV-2 variants in three ancestry groups. **Legend**: **A**. WHICAP cohort ancestry distribution of all common and rare variants in the *LPA* KIV-2. **B.** previously identified (published) exonic variants in the *LPA* KIV-2, **C.** WHICAP novel exonic variants, **D**.WHICAP novel intronic variants. NHW: Non-Hispanic White, B: Black, CH: Caribbean Hispanics. There are no previously published studies identifying intronic variants in this region.

A large proportion of the variants (n=1007, 70.9%), were rare (the minor allele was observed in less than 1% of the variants). The data for novel and previously identified exonic variants, including sex and ancestry distribution, are presented in Supplemental Figure S2.

### Association of *LPA* KIV-2 variants and KIV-2 copy number with WHICAP metabolic risk factors

Ten of our *LPA* KIV-2 variants in the WHICAP cohort were significantly associated with our WHICAP metabolic risk factors **(Table 2**, and **Figure 2)**. Three novel exonic, common variants, were present in all ancestry groups and showed significant positive associations with HTN; 4785-C/A [overall carrier frequency (CF) = 78%, OR=1.45, p=0.032], 727-T/C (CF=96%, OR=2.11,p=0.032) and 723-A/G (CF=96%, OR=1.97, p=0.038). Six intronic variants were also positively associated with HTN (166-G/A, 387-G/C, 402-G/A, 4527-A/T, 4541-G/A, 4653-A/T). One additional intronic variants, 412-C/T (CF=76%, β= −14.52, p=0.02) was negatively associated with blood glucose level and the only significantly associated variant not associated to KIV-2 CN. The CF of most variants had broad ranges within each ancestry group (**Supplemental Table S1**), however, the CF of these 10 variants are generally consistent and mostly common ( CF between .03 to .97) across the three ancestry groups, **Table 2**, and **Supplemental Table S3A.** Among the nine variants highly associated with HTN, we observed modest to high linkage disequilibrium (LD) between 723-A/G and 727-T/C (D’ = 0.846, r^2^ = 0.655), 4527-A/T and 4541-G/A (D’ = 0.787, r^2^ = 0.49), 387-T/C and 402-G/A (D’= 0.923, r^2^ = 0.283), **Supplemental Table S3D.**

**Figure 2.**
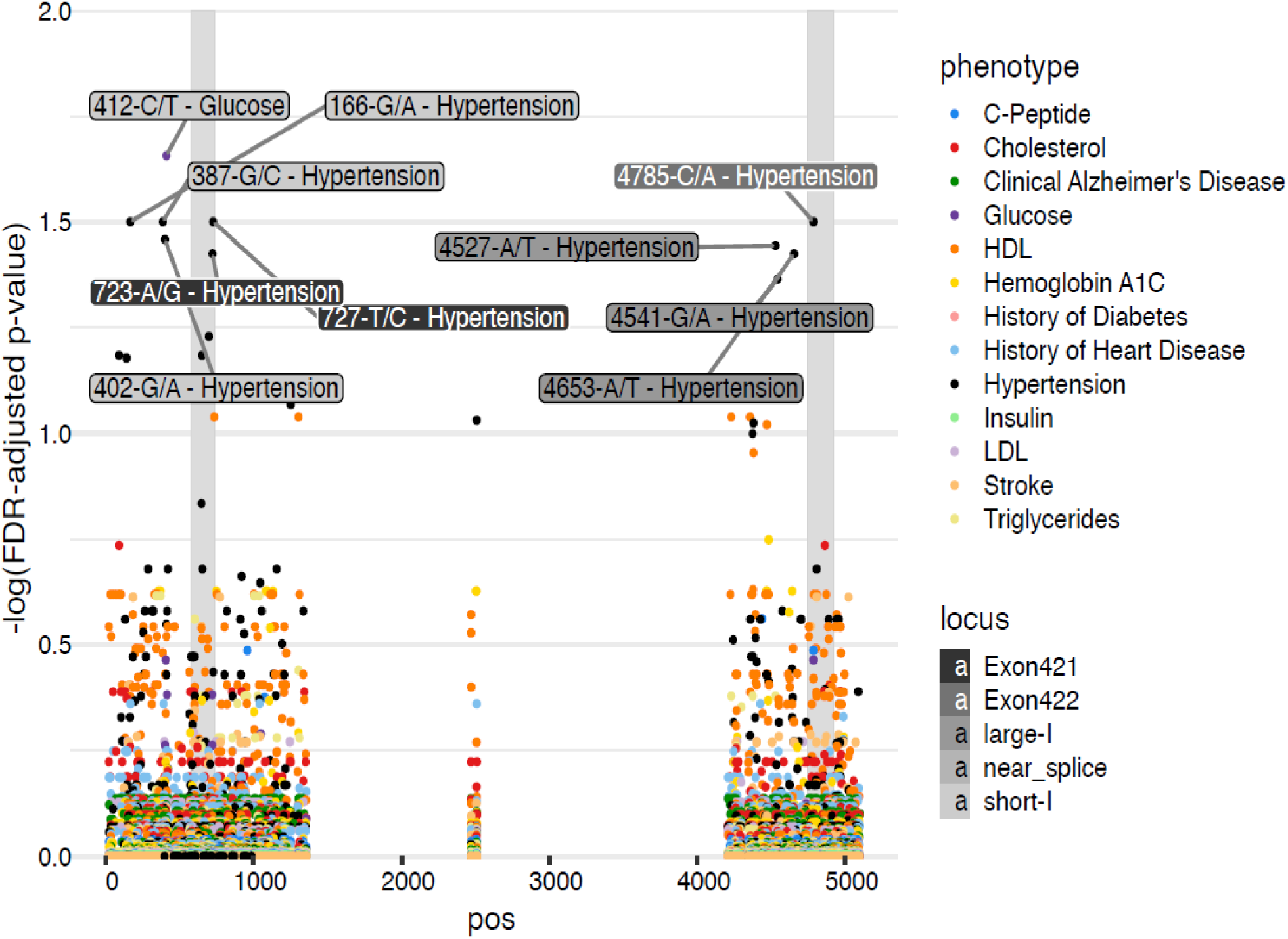
Association of *LPA* KIV-2 variants and KIV-2 CN with WHICAP metabolic risk factors. **Legend:** Association plot showing the distribution and association of *LPA* KIV-2 variants with WHICAP phenotypes. X-axis-Pos: gene locus relative position. Y-axis: false discovery rate adjusted p-value.

**Table 2.**
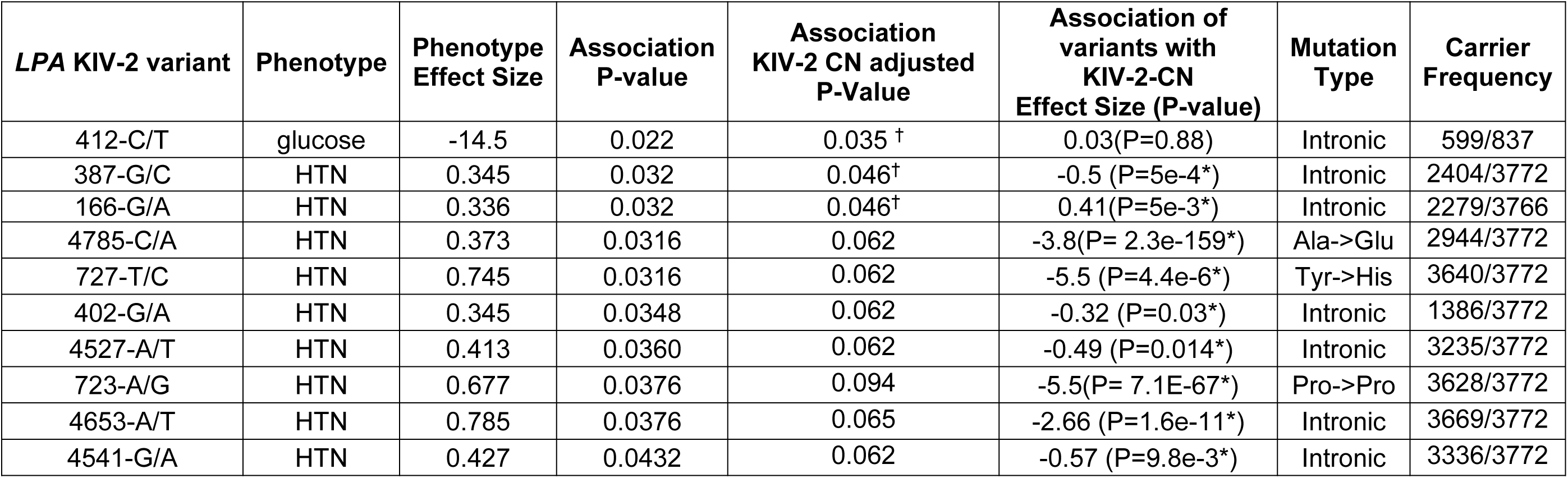
Association of novel *LPA* KIV-2 variants with glucose levels and hypertension. **Legend:** Associations for glucose were testing using OLS and logistic regression was used for HTN. The effect sizes for all associations were in the same direction (+ or -) for the three ancestry groups (NHW, B, CH). P-value are all FDR adjusted and only significant variants are listed on the table. Full list of the associations of variants with KIV-2 CN are listed in supplementary table S4. (+): positive associations, (-): negative association. ^†^Variant association with phenotype remained significant after adjusting for KIV-2 CN; *variant with significant association with KIV-2 CN NHW: non-Hispanic white, B:Black, CH: Caribbean Hispanic; Kringle IV type 2, Glucose: blood glucose levels, HTN – history of hypertension, CN: copy number; NA: not applicable.

There were a total of 314 variants that were significantly associated with KIV-2 CN (**Supplemental Table S4**) and the KIV-2 CN was negatively associated with plasma cholesterol levels an overall negative trend with LDL-C levels. These associations were driven by the NHW population and may be related to the contamination of LDL-C measurements and Lp(a), which has been previously described^31^ **(Supplemental Table S5)**. Three of the above intronic variant associations [412-C/T (β=-14.2, p=0.03) (glucose),166-G/A (OR=1.41, p=0.05) (HTN), 387-G/C (OR=1.40, p=0.05) remained significant when taking KIV-2 CN into account, among these 3 variants, only two 166-G/A and 387-G/C are independently associated with KIV-2 CN **(Table 2)**.

### Association of *LPA* KIV-2 variants with KIV-2 copy number

The ancestry-wide and population-overall distribution of KIV-2 CN estimate can be seen on **Figure 3 (A, B, C).** Not surprisingly, as Lp(a) levels are highly regulated by this region, a large number of variants were significantly associated with KIV-2 CN. The majority of the significant associated variants (206 of 314, 65.6%) were related to a reduction in the number of KIV-2 repeats (CN) [i.e. higher levels of Lp(a)] e.g. 4894-G/A (beta = −4.01, FDR-adjusted p = 1.17e-284), 690-A/C (beta = −3.95, FDR-adjusted p = 2.48e-273), **Figure 4**. While 34.5% percent were correlated to an increase in the CN [i.e. lower levels of Lp(a)].

**Figure 3.**
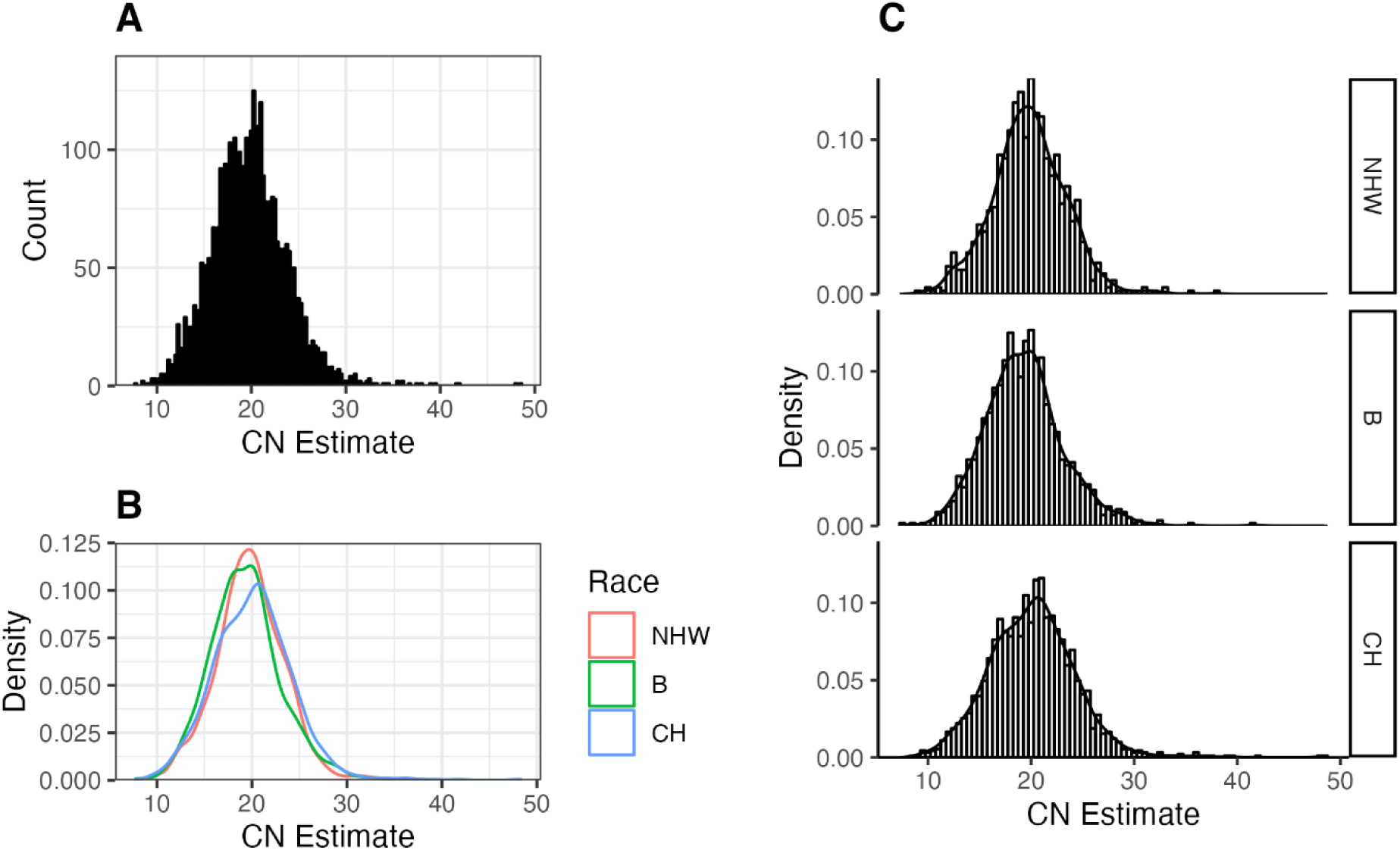
*LPA* KIV-2 copy number distribution in three ancestry groups. **Legend: A**. Overall Distribution of *LPA* KIV-2-CN in WHICAP cohort. **B/C.** Ancestry specific *LPA* KIV-2 CN distribution in WHICAP cohort. NHW: Non-Hispanic White; B: Black individuals; CH: Caribbean Hispanics. Density: probability density function.

**Figure 4:**
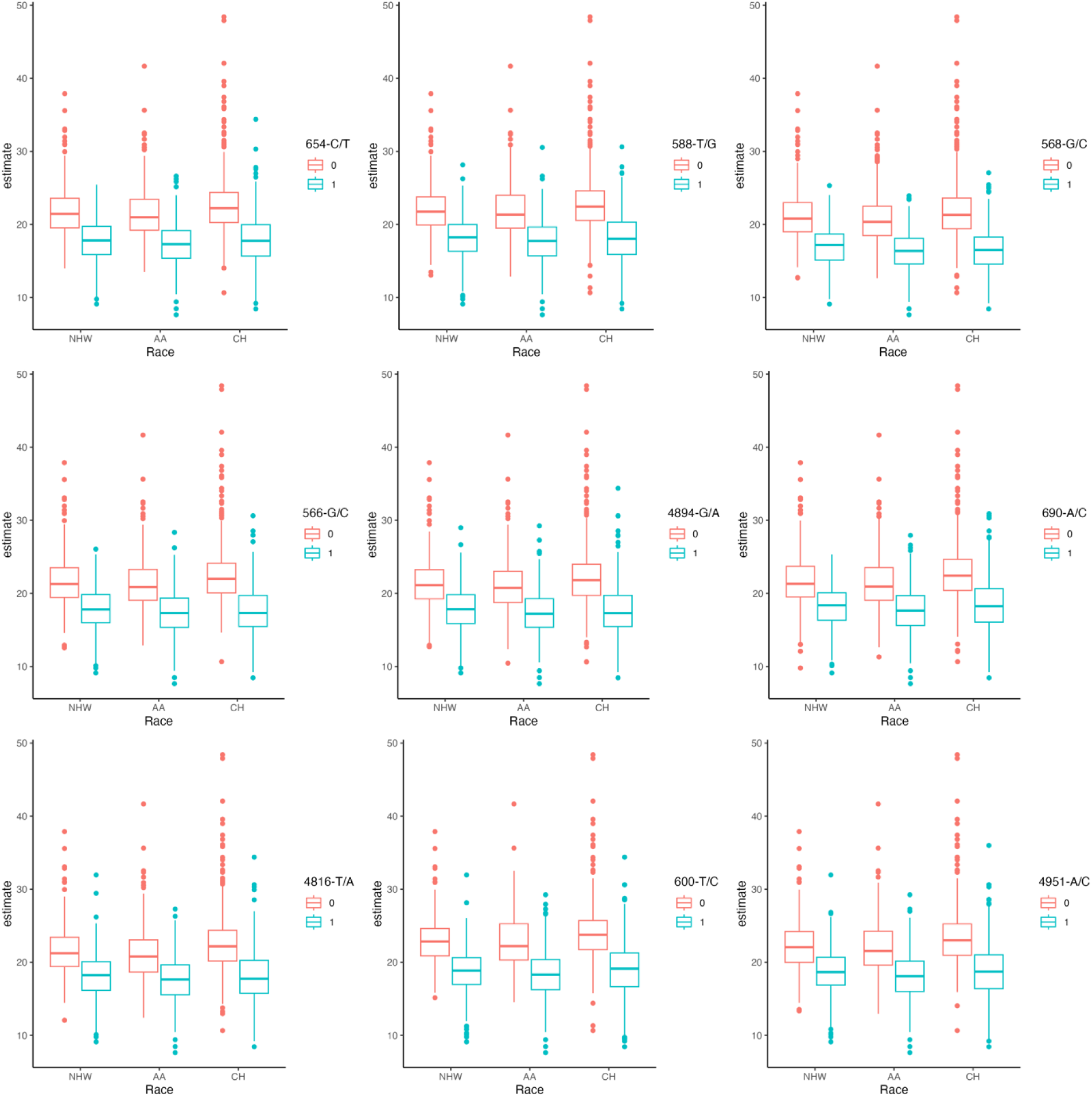
Top nine *LPA*-KIV-2 variants significantly associated with the number of KIV-2 repeats. **Legend:** Box-plots of top nine LPA KIV-2 variants affecting KIV-2 copy number. NHW: non-Hispanic Whites, B: Blacks, CH: Caribbean Hispanics.

The WHICAP cohort has been extensively examined for genetic risk of AD and APOE-ε4 genotype have been tightly linked to AD and other related traits^23, 32^. Since APOE-ε4 is a strong genetic risk factor for AD and cardiovascular phenotypes, we controlled for the dosage of APOE-ε4 allele in all our analysis. The distribution of APOE-ε4 allele in our population was: non-carriers (ε33: 2240, ε23:490 and ε22:25)- 2755 (B 718, NHW 681, H 1356), ε4-heterozygotes(ε24:104, ε34:854 ) - 958 (B 357, NHW, 185 H 416), and ε4 homozygotes-59 (B 29, NHW 13, H 17), 2 individuals have their information missing in the dataset.

### Association of LPA KIV-2 variants with plasma Lp(a) levels and apo(a) isoforms: Pilot Study

In a small number of serum samples (N= 27; CH: N=14, B: N= 8, NHW: N= 5) from the WHICAP cohort (**Supplemental Table S6,** Baseline Characteristics), we evaluated the association of Lp(a) levels and isoform size (i.e. weighted isoform size) with LPA KIV-2 variants, controlling for APOE-ε4 genotype and KIV-2 CN (**Supplementary Table S7**, Individual data for 27 subjects). The median Lp(a) levels for this population were relatively low 18.87 nmol/L.As expected, there was a strong association between our estimated isoform size (wIS) ( ß=0.74. 5e-3) and KIV-2 CN. Additionally, the wIS was negatively associated with Lp(a) levels.

In this small cohort, 107 variants within the *LPA* KIV-2 region met our criteria for selection, **Supplemental Table S8A and B.** Twenty-five of the variants are exonic, of which 9 have been previously reported^19^ and 16 are novel to the data set. We found 1 significant association between *wIS* and previously reported intronic variant (937-T/C) with β = −8.05, FDR-adjusted p=0.02. However, this association is lost when we correct for race in this small cohort. Two exonic variants 4816-T/A (p=0.002) and 4783-G/C (p=0.002)showed a positive association with plasma Lp(a) levels and negative association with KIV-2 CN. These variants were not in LD. The direction of these associations is consistent with the expectation that *wIS* is positively associated with KIV-2 CN and negatively associated with Lp(a) level, **Table 3**. (**Supplemental Table S8**).

**Table 3:** Association of *LPA* KIV-2 variants with weighted isoform size and Lp(a) levels in pilot cohort. Legend: Only significant associations are listed on this table, raw data presented on supplemental **Table S4**, **S8A** and **S8B**. Lp(a): Lipoprotein (a); *wIS*: weighted isoform size; CF: carrier frequency; Beta: effect size, p-values: calculated from linear regression. As Lp(a) levels are not normally distributed we log transformed for obtaining the beta for Lp(a).

| Variant | wIS<br>p-value | Lp(a) levels<br>p-value | KIV- 2 CN<br>p-value | Carrier<br>Frequency | Mutation<br>Type |
| --- | --- | --- | --- | --- | --- |
| 4908-A/G | NS | 0.01 | 1.6E-232 | 7/27 | Exon |
| 4894-G/A | NS | 0.013 | 1.2E-284 | 7/27 | Exon |
| 4816-T/A | NS | 0.027 | 6E-268 | 11/27 | Exon |
| 520-G/T | NS | 0.034 | 2.27E-46 | 16/27 | Intron |
| 937-T/C | 0.02 | NS | 1.4E-193 | 13/27 | Intron |

We wanted to understand the role of these 3 variants in the larger (WHICAP) cohort and if any were associated with metabolic phenotypes. Indeed, the two exonic variants positively associated with Lp(a) levels were negatively associated with KIV-2CN. However, there were no significant relationships of these variants with clinical phenotypes (Supplemental Table S3B, all associations non-significant, p>0.05).

## Discussion

On a per particle basis, apo(a), is thought to be more atherogenic than apoB100, the main carrier protein of low density lipoprotein (LDL)^33^. The binding of apo(a) particles to liver or plasma circulating apoB100 particles allows the formation of Lp(a) whose levels are highly regulated by the *LPA* gene^34^. Lp(a) levels are higher in Blacks and Hispanics^35, 36^, even at similar isoform size ^10, 37, 38^ and recent data from the multi-racial Lp(a) HERITAGE study^36^ showed that median Lp(a) levels were higher in Black patients with ASCVD (132 nmol/L, [∼53 mg/dL]) compared to the overall group (52 nmol/L, [∼21 mg/dL]). The HERITAGE study had limitations in the assays used to measure Lp(a) levels however, this data has been reported by others in other multiethnic cohorts ^39^;^40^ The pathophysiology relating to these higher absolute levels of Lp(a) in Black individuals remain unknown. Lastly, standardization and harmonization of Lp(a) plasma measurement are still in development, hence using genetic data to link to disease may provide insights beyond circulating plasma Lp(a) levels.

Our current study finds that the LPA KIV-2 region contains many variants that are expressed in all ancestries, albeit some expected differences in carrier frequencies. Recent exploration of the highly repetitive *LPA* KIV-2 region highlighted various single nucleotide variants and mutations ^8^, however the data was limited in racial diversity that is represented in our current study. This region has strong regulations of Lp(a) levels and therefore understanding its variance could help with identifying those that are at increased risk of disease. Moreover, the relationships of LPA KIV-2 variants to cardiovascular and other metabolic outcomes have not been previously studied. The latter of high importance as we try to understand why there are racial differences in the levels of Lp(a) and if these are linked to disease presentation.

Our positive associaitons linking unique LPA KIV-2 variants are insightful as published studies have reported high Lp(a) concentrations in hypertensive individuals when compare to normotensive subjects, however these data are not consistent ^41–45^. The differences in these associations could be due to our current study findings of the multiple genetic variants that could be linked to Lp(a) and HTN. Additionally, identifying the unifying pathological pathway that links high Lp(a) levels to HTN would be an important area of future research and has been recently reviewed by others^46^. Interestingly, recent evidence from the MESA cohort, another multi-ethnic cohort, revealed that high Lp(a) levels enhanced the association of hypertension with incident CVD, and this association was the greatest among Black participants ^43^. The latter highlighted in our data showing that association p-values (for variants and links to HTN) were lowest in those of B ancestry compared to CH and Whites. Additionally, a recent retrospective analysis of large cohort, showed that elevated Lp(a) levels were associated with a higher ASCVD risk in males, hypertensive, and diabetic patients^47^.

There are various studies that have linked Lp(a) to incidence of T2DM, however, there is no conclusive data with mechanism or exact pathology driving this observation^48 15^. The negative association of one of our variants with glucose levels could explain some of this observed phenotype. Importantly, this novel variant association with glucose levels was not affected by the KIV-2 CN. Previous studies have reported low levels of Lp(a) have been associated with T2DM ^15, 49, 50^. In a Danish cohort the highest quintile of the sum of the KIV-2 repeats from the two apo(a) alleles was associated with T2DM. These reports, including our variant association with glucose levels will need to be validated and in specific the links of why low Lp(a) levels are link to a potential insulin resistance pathways that controls T2DM. There are non-LPA KIV-2 variants that have been identified in large populations and may play a role in Lp(a)-related diseases such as rs3798220 ^51–53^. Rs3798220 is infrequent in individuals of European (non-Hispanic Whites-NHW) and AA ancestry [carrier frequency (CF)=0.03,0.02, respectively] but common in Caribbean Hispanics(CH), (CF=0.12). Rs3798220 is a coding variant located in the protease-like domain of *LPA* and is associated with plasma Lp(a) levels and cardiovascular risk, but not in all populations^54^. Further, in the WHICAP cohort, rs3798220 was associated with baseline insulin c-Peptide levels (beta=3.4, Wilcoxon p-value=0.007), with variant carriers having higher c-peptide levels. This data may support recent observed association of hepatic insulin resistance with Lp(a)^55^.

The current data set highlights the various genetic influences that may be driving disease presentation in those with high Lp(a) levels. It is well established that a dysfunctional endothelium^56–58^ is at the core of HTN disease, the latter may also be linked to diabetes. In vitro studies have shown that elevated Lp(a) can aid in the process of atherogenesis and cause endothelial dysfunction, mainly driven by the apo(a) component^59^ of the particle which is the product of the *LPA* gene.

Importantly, previous reports showed that two splice site variants, 4925G>A and 4733 G>A, were highly frequent (22% and 38% are carriers, respectively) and result in a pronounced decrease in Lp(a) concentration (by ≈30 and 13 mg/dL, respectively) and a lower risk for cardiovascular disease. These two variants are the second and third strongest relative contributors to Lp(a) concentration beyond apo(a) isoform size. Yet, these variants are not present in our WHICAP cohort and highlight the need to study diverse cohorts to understand population drivers of Lp(a) levels and disease.

Marcovina et al. showed that for a given apo[a] size, Blacks commonly have higher plasma levels than do Caucasians^60–62^. Even within a racial group, there is considerable variation in levels for a given apo[a] size, suggesting additional regulatory factors. Our study supports these findings by the discovery of the specific variants present in Black individuals and CH that are not observed in White population. Moreover, is important to note that in our study the variants that were significantly linked to outcomes were present in all ancestry groups, suggesting that CF may be more important moving forward.

Enkhmaa et al. ^10^ examined a large population of NHW and Blacks and noted some racial differences in allele presentation between the races. Heterozygotes with at least one detectable apo[a] isoform (238 Caucasians and 159 African Americans), had various differences. In the current study, when we examined *LPA* KIV-2 CN we observed that only four of our significant variants were affected by the KIV-2 CN. It is important to note that in the previous studies there were no interactions between apo[a] alleles within genotypes; apo(a) specific isoform levels were not affected by the difference in size. Additionally, the authors in that study noted that the dominance pattern was explained by Lp[a] level and apo[a] genotype in Blacks more than in Caucasians (29% vs. 13%).

In a small cohort of WHICAP participants, we were able to measure Lp(a) levels and isoform size. We identified 3 novel *LPA* KIV-2 variants that were associated (two positively associated and one negatively associated) with isoform size and one negatively associated with weighted isoform size (Table 3). These novel associations will need to be validated in larger cohorts that have both Lp(a) serum levels and genetic level data sets including apo(a) isoforms.

Our current findings have to be reviewed in the context that a genetic variant can be in strong linkage disequilibrium with the real causal variant and without functional studies it will be hard to determine which variant is functionally responsible for Lp(a) concentration variability. It is unclear how these variants affect the production or clearance of apo(a) or the proteins that bind to the particle. Lp(a) transcription may be affected by these variants and some of the regulators of Lp(a) transcription have been recently reviewed^63, 64^.

In addition, following recently published studies where investigators created haplotypes to increase the number of SNVs (instead of single SNV for risk prediction) followed by a GWAS for myocardial infarction^65^, opens up additional work that can be derived from our studies. Others have used qPCR to quantify the copy number of KIV type-2 repeats in large cohorts from the Copenhagen study. This method sums up the number of both apo(a) alleles and cannot distinguish each of the two alleles^9^.

The current study findings of novel variants and their association with cardiovascular disease and other risk factors expands our knowledge significantly. As the KIV-2 region is one of the main drivers of circulating Lp(a) levels, these newly identified relationships can assist us with understanding disease mechanisms. The findings also highlight the unique common and rare variants that exist based on ancestry.

### Study Limitations

We used exome-wide reads to re-align to the KIV-2 reference^8^, to ensure all unaligned reads with potential matches to the repeat are aligned in the resulting BAM file. It is conceivable that reads aligning to regions of high similarity in other Kringle repeats could result in false positive variant calls. To partially overcome this limitation, we only used variants observed in 5 or more individuals within each ancestry group, assuming that recurrent variants are likely true calls. Our study concentrated on the analysis of variants within the *LPA* KIV-2 regions, however, there is still a wide variability in Lp(a) concentrations within each KIV-2 repeat group that could be explained by non-genetic factors and other genetic variants than the KIV-2 repeats. Several of these variants within the *LPA* gene region have been described and have been linked to higher levels of Lp(a), mostly in Caucasian populations^66^. The data on diverse racial groups is not as strong and hence our study adds to the field on those efforts. Differences in carrier frequency and prevalence of cardiovascular risk factors between and within ancestral groups could be confounders and result in false positive association. To overcome this, we analyzed each ancestral group independently and including PCs that capture latent population substructure to account for carrier frequency differences. Importantly, the disease relevance of these variants and their links to KIV-2 CN have not been previously studied. Our pilot data cohort, which had Lp(a) plasma measurement, is small and hence the effect sizes and correlations with Lp(a) levels in this small cohort may not extrapolate to larger populations. However, the relationships with KIV-2 were also present in the larger cohort.

## Conclusion

We have identified novel *LPA* KIV-2 variants in three ancestry groups and report novel associations to HTN, glucose and KIV-2 CN. Additional validation studies using available diverse data sets with access to outcome data will be needed to confirm the link of novel variants to Lp(a) levels and disease presentation. Additionally, the pathways regulating these associations should be further characterized.

## Data Availability

All the data is available on a server and by request from study senior author.

https://github.com/alexliyihao/vntrwrap

https://github.com/alexliyihao/lpa-analysis

## Acknowledgments

The authors will like to thank the WHICAP participants. The Lp(a) plasma levels and isoform size were measured by Medpace, Inc. under the expertise of Dr. Santica Marcovina. We than Dr. Adam Bress, Associate Professor of Population Health Sciences in the Division of Health System Innovation and Research, University of Utah Schoool of Medicince for expert discussions surrounding our variant associations with hypertension.

## Supplemental Material

**Table S1.** Full data set for WHICAP variant calling (*excel file*).

**Table S2.** Number of WHICAP *LPA* KIV2 Variants and appearance frequencies compared to 1000 Genome Cohort.

**Table S3A:** Ancestry carrier frequencies for WHICAP variants with significant associations to metabolic risk factors

**Table S3B:** Association of *LPA* KIV-2 variant vs. metabolic risk factors in WHICAP (excel file) **Table S3C:** Association of *LPA* KIV-2 variant vs metabolic risk factors in WHICAP, adjusted by *LPA* KIV-2 CN (excel file)

**Table S3D** Linkage Disequilibrium between *LPA* KIV-2 variants significantly associated with hypertension (excel file)

**Table S4:** Association of *LPA* KIV-2 CN vs. *LPA* KIV-2 variants in WHICAP (excel file)

**Table S5:** Association of *LPA* KIV-2 CN with metabolic risk factors

**Table S6.** Baseline Characteristics: Pilot validation study

**Table S7.** Individual characteristic pilot cohort

**Table S8.** *LPA* KIV2 variant association with weighted isoform size (**S8A**) and Lp(a) serum levels (**S8B**) in Pilot Study individuals (N=27) (excel file).

**Figure S1.** WHICAP *LPA* KIV-2 Rare and Common Variants

**Figure S2.** Sex and ancestry distribution of exonic and intronic variants.

## Non-standard Abbreviations and Acronyms

WHICAP: Washington Heights Inwood Community Aging Project
Lp(a): Lipoprotein (a)
wIS: weighted Isoform Size
ASCVD: Atherosclerotic Cardiovascular Disease

